# Safety and Efficacy of Preventative COVID Vaccines: The StopCoV Study

**DOI:** 10.1101/2022.02.09.22270734

**Authors:** Sharon Walmsley, Leah Szadkowski, Bradly Wouters, Rosemarie Clarke, Karen Colwill, Paula Rochon, Michael Brudno, Rizani Ravindran, Janet Raboud, Allison McGeer, Amit Oza, Christopher Graham, Amanda Silva, Dorin Manase, Laura Parente, Jacqueline Simpson, Roaya Monica Dayam, Adrian Pasculescu, Anne-Claude Gingras

**Author notes:** **Corresponding author:**, 416-3340-3871.

## Abstract

**Background:** To partially immunize more persons against COVID-19 during a time of limited vaccine availability, Canadian public health officials recommended extending the vaccine dose interval and brand mixing. Impact on the antibody response among the older ambulatory population was unclear.

**Methods:** Decentralized prospective cohort study with self-report of adverse events and collection of dried blood spots. Data is presented for 1193 (93%) of the 911 older (aged >70 years) and 375 younger (30-50 years) recruits.

**Findings:** Local and systemic reactivity rates were high but short-lived, particularly in the younger cohort and with mRNA-1273 vaccine. After a single COVID-19 vaccine, 84% younger but only 46% older participants had positive IgG antibodies to both spike protein and receptor binding domain (RBD) antigens, increasing to 100/98% with the second dose respectively. In multivariable linear regression model, lower normalized IgG RBD antibody ratios two weeks after the second dose were statistically associated with older age, male gender, cancer diagnosis, lower body weight, BNT162b2 relative to mRNA-1273 and longer dose intervals. Antibody ratios in both cohorts declined 12 weeks post second vaccine dose.

**Interpretation:** We report success of a decentralized serology study. Antibody responses were higher in the younger than older cohort and were greater for those with at least one mRNA-1273 dose. The immunity threshold is unknown but correlations between binding and neutralizing antibodies are strongly positive. Trends with time and at breakthrough infection will inform vaccine booster strategies.

**Funding:** Supported by the Public Health Agency of Canada and the University Health Network Foundation.

## Introduction

Clinical trials and population-based studies demonstrate excellent short term efficacy and safety profiles for mRNA vaccines. ^[1-4]^ Few people over age 70 were included in the randomized vaccine trials^[1]^, yet older persons, especially those with comorbidity, have higher risk of mortality from COVID-19 infection. Natural antibody appears protective against re-infection ^[5, 6]^ but we currently are unable to determine the immunity threshold COVID-19 vaccines confer against asymptomatic infection and transmission^[7]^ or against variant strains.^[8]^ Longitudinal vaccine antibody response data outside of the clinical trial setting and at the time of breakthrough infection will help inform the priorities and timing of booster doses.

To partly immunize more Canadian population at a time of supply issues, the National Advisory Committee on Immunization recommended extending the dose interval up to 4 months and allowed vaccine brand mixing. These recommendations raised concern about vaccine efficacy. While some studies retrospectively reported advantageous immune responses from extended doses, the consequences in the older population is unknown.^[9]^

Our primary objective was to compare the safety and antibody responses to COVID-19 vaccines in an older community dwelling cohort relative to a younger cohort. We hypothesized the older group would have a less robust response.

## Methods/Design

### Design

A decentralized longitudinal cohort study planned to follow participants with two COVID-19 vaccine doses for 48 weeks and participants with three doses for 96 weeks. We report results to 12 weeks post the second vaccine dose. The full protocol is available on the study website www.stopcov.ca. Trial registration: Clinicaltrials.gov. NCT05208983.

### Recruitment

A data sharing agreement with the Ontario Ministry of Health enabled us to send study information emails to persons receiving the COVID-19 vaccine at an Ontario distribution center who consented for contact for research. A similar email was sent to Ontario Canadian Association of Retired Persons members (www.carp.ca). Participants could enrol through the website prior to the first or second vaccine dose.

### Electronic consent

including the request to share core data elements with the Canadian Immunity Task Force was completed on the study website. A video and periodic questions were added to enhance comprehension. The study and electronic consent process were approved by the University Health Network (UHN) Ethics Review Committee. Consented participants used the study website with their personal identification (ID) number and password as a portal for communication with study staff, data collection and results reporting. A schedule for required activities and email reminders are provided.

### Questionnaires

Self-administered electronic questionnaires collected baseline demographic and health data. Electronic diaries collected data on vaccine date and brand as well as local and systemic adverse events for 7-days after each vaccine dose. Monthly check-in questionnaires capture persistent adverse events, booster doses and new COVID-19 diagnoses.

### Dried Blood Spot (DBS) Specimens

Protocols for self-collection and shipping of DBS specimens were adapted from those previously shown feasible^[10]^. A commercial company prepared and distributed the kits. Instructions were provided in hard copy and in video on the website. Samples were requested +/-7 days of initial vaccine, three weeks (+/-1week) after the first vaccine dose, two weeks (+/-1week) after the second vaccine dose and then every 12 weeks. If the dose interval exceeded 28 days, an additional sample was collected prior to the second vaccine dose. DBS were collected on Whatman 903 cards using a lancet for finger-prick.

### Laboratory Methods

### Serological Assays

Completed DBS cards were mailed to the UHN research unit, checked for quality, logged and transferred to the Lunenfeld-Tanenbaum Research Institute (LTRI) for processing. Antibodies were eluted from punches from the DBS as previously described and tested by Enzyme Linked Immunosorbent Assay (ELISA) for antibodies (IgG) against the spike trimer, its receptor binding domain (RBD) and nucleocapsid proteins (NP) ^[11, 12]^. As samples with high antibody levels saturate the assays preventing accurate measurement, all samples were tested at two dilutions (2·5ul of eluate/well (1:4) and 0.156 ul/well (1:64)) to ensure a large fraction of the measures would be tested within the linear range of quantification^[12]^. We selected to profile total IgG antibodies to the indicated antigens since our results and others ^[11]^ show a strong correlation especially between anti-RBD IgG levels and neutralization titers, enabling us to infer neutralization changes across groups. The assay was validated by duplicate Receiver Operating Characteristics analysis of positive PCR confirmed cases and negative pre-COVID samples donated by the National Microbiology laboratory of Canada.^[12]^

### Interpretation and Reporting of Results

Results are reported as relative ratios to a synthetic standard included as a calibration curve on each assay plate^[12]^. For the standard curve we used recombinant antibodies against spike and RBD. Seropositivity for each assay is defined by calculating the mean +/-3 standard deviations (SD) in aggregate data from negative controls. Seropositivity cutoffs are normalized ratios of 0·482 for anti-spike IgG, 0·324 for anti-RBD IgG and 0·642 for NP using the highest concentration (2·5 ul/well). Specimen overall positivity to vaccine required the IgG’s to both spike and RBD to be above threshold. Relative antibody ratios were also compared with the median levels of convalescent serum obtained 21-115 days after symptom onset in patients with COVID-19. Calibration of these assays to WHO standard was previously described. ^[12]^

### Data management and Statistical Analysis

#### Sample Size Considerations

A sample size of 768 older and 192 younger participants was planned to enable detection of differences in normalized ratios of anti-spike and anti-RBD IgG antibodies of ·092 and ·124 at two weeks post second vaccination, with 80% power and a significance level of ·05, assuming SDs of ·406 and ·55 respectively^[11]^. We anticipated 25% dropout post second vaccination (either through attrition or through poor quality DBS) and adjusted our target sample size to a total of 1286.

### Statistical Analysis

Baseline characteristics, vaccine type, and time between doses were compared by age group using chi-square, Fisher’s exact, Cochran-Armitage, or Wilcoxon rank-sum tests as appropriate. Antibody ratios were compared between age groups, vaccine type using chi-square, Fisher’s Exact, Kruskal-Wallis and Wilcoxon rank-sum tests. Pearson correlation coefficients were used to describe the association between vaccine interval and antibody ratios. Univariable and multivariable linear regression was used to model antibody ratios to RBD at two and twelve weeks (separately) after the second vaccine dose and test for an association with age. Covariates included in the multivariable models were chosen a priori based on clinical expertise and included gender, comorbidities (cardiovascular disease, cancer, diabetes, immunosuppression), body mass index (BMI), vaccine type, and time between doses.

## Results

A total of 1286 adults (911 older and 375 younger) were recruited between May 17-July 31, 2021. Five participants did not meet eligibility criteria, 20 withdrew consent and 68 were lost to follow-up leaving 1193 (93%) in the current analysis. Fifteen participants (14 aged 30-50 years) were recruited prior to their first vaccine dose, the remaining 1178 prior to their second vaccine dose reflecting the timelines of study initiation and vaccine distribution.

The baseline questionnaire was completed by 1186 (99%) participants. Table 1 summarizes baseline characteristics stratified by age group. 60% of the older and 76% of the younger cohort were female or non-binary. A greater proportion of the older cohort were white (93% vs.75%) and more overweight or obese compared to the younger cohort. The older cohort had more comorbidity.

**Table 1:**
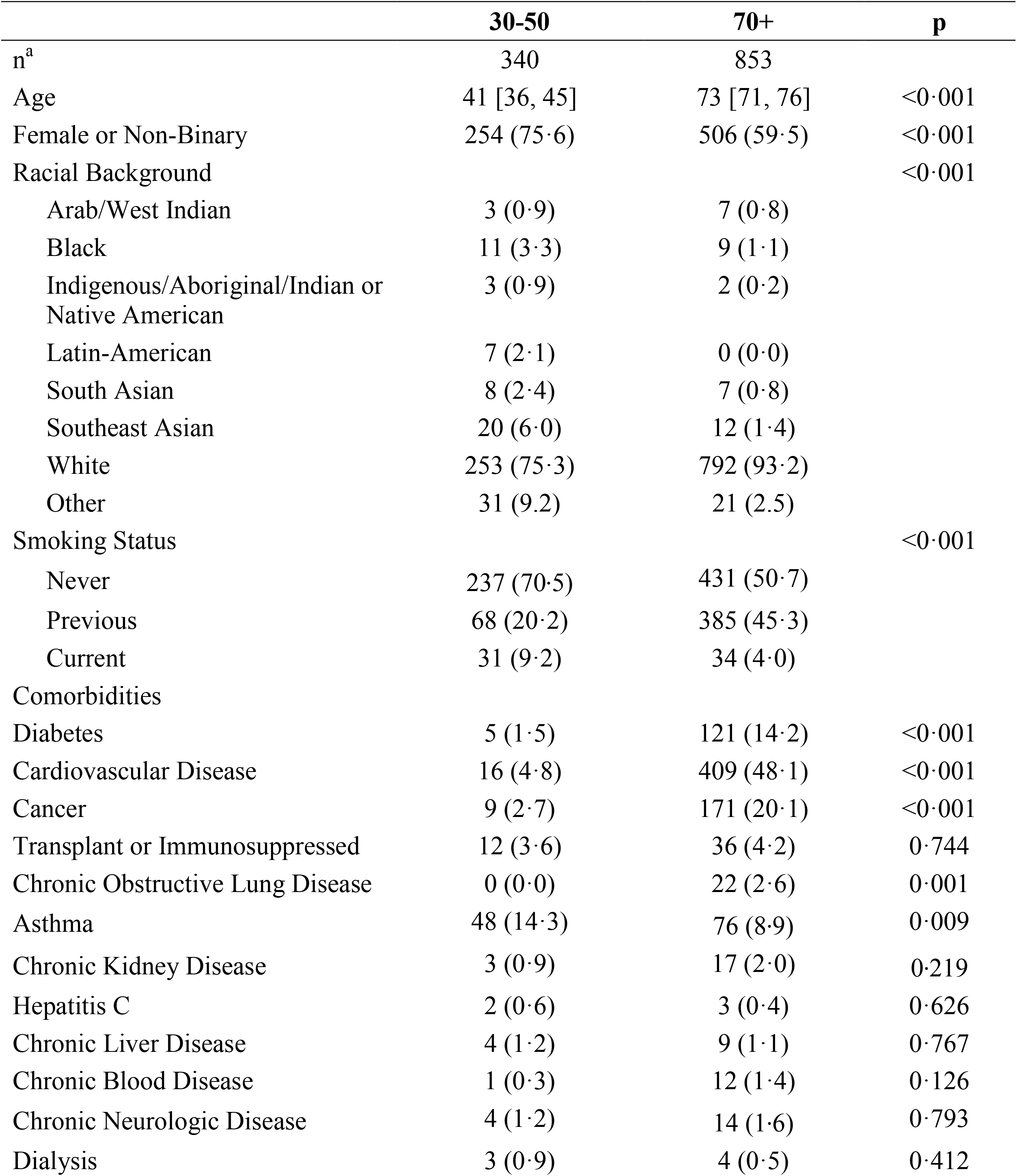
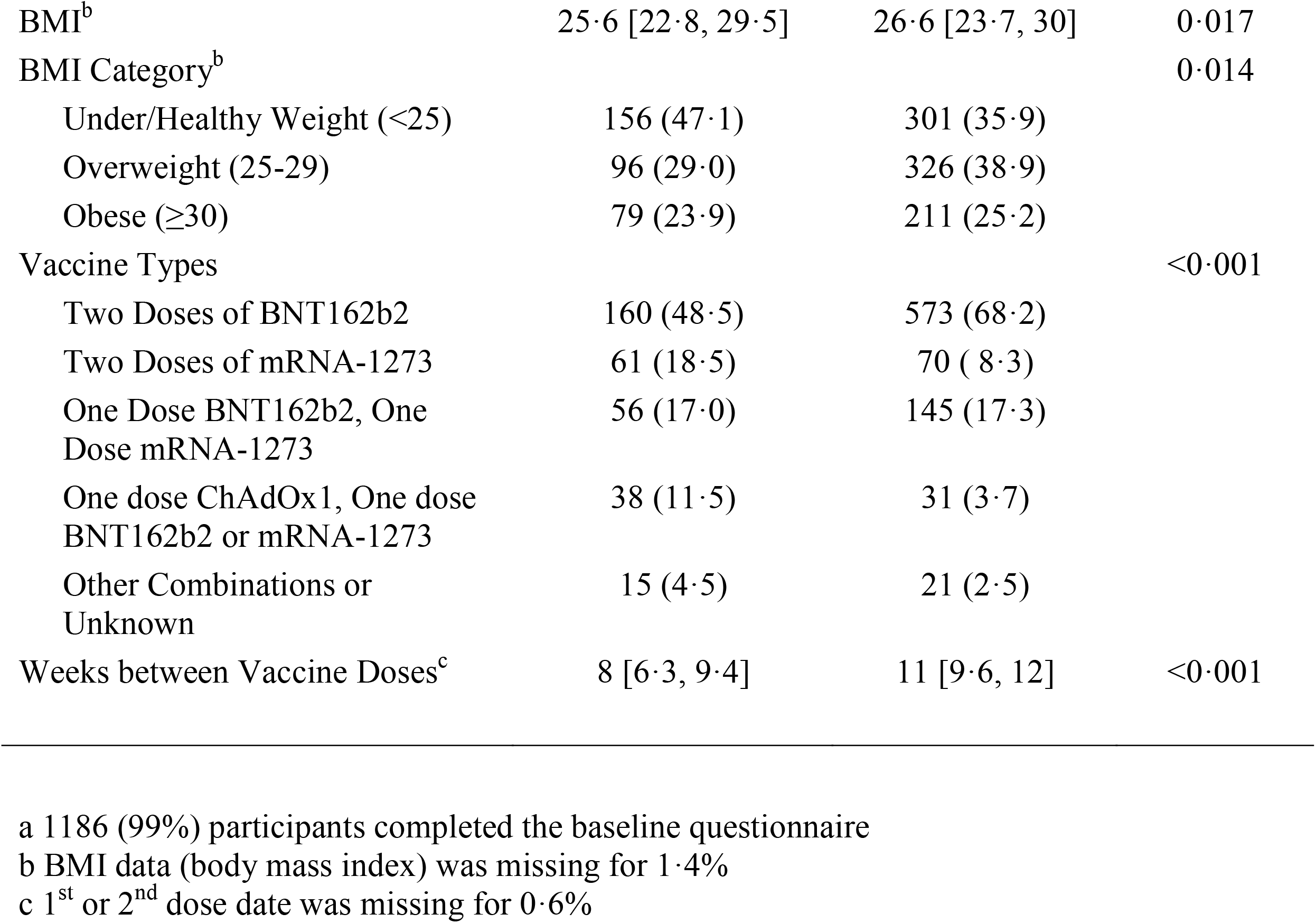
Baseline Characteristics and COVID-19 Vaccine Brands by Age Cohort

Most participants received two doses of either BNT162b2 (Pfizer-BioNTech) or mRNA-1273 (Moderna) with older participants more likely to receive BNT162b2 (68% vs. 49%) and younger participants more likely to receive mRNA-1273 (19% vs. 8%). 17% in both cohorts received one dose of each brand of mRNA vaccine. 11·5% of the younger group and 4% of the older group received one dose of ChAdOx1 (Astra Zeneca) and one dose of an mRNA vaccine. The interval between vaccine doses was a median (IQR) of 11 (9·6, 12) weeks for the older cohort and 8 (6·3, 9·4) weeks for the younger cohort.

### Safety

The 7-day diaries were completed for 37 (3%) participants after the first vaccine dose and 955 (80%) after the second vaccine dose. After the first dose, the most commonly reported adverse events were pain near the injection site (70%), fatigue (60%), and malaise (46%). Adverse events reported after the second vaccine dose are presented in Figure 1. The most commonly reported events were pain near the injection site, fatigue, muscle aches or pains, malaise, and headaches. Younger participants were more likely to experience each adverse events compared to the older participants (Figure 1a), and also more likely to experience at least one event to a moderate (some interference with activity) or severe (significant, prevents daily activity) degree (82%) compared to the older participants (39%). Adverse events after the second dose were more likely for participants receiving mRNA-1273 compared to BNT162b2 (Figure 1b). At seven days (Figure 1c), few participants reported symptoms. Overall 49 (5%) participants reported no adverse events to the second vaccine dose. At the monthly check-in, 4% reported persistent adverse events thought related to the vaccine at month 1, decreasing to 1% at month five.

**Figure 1:**
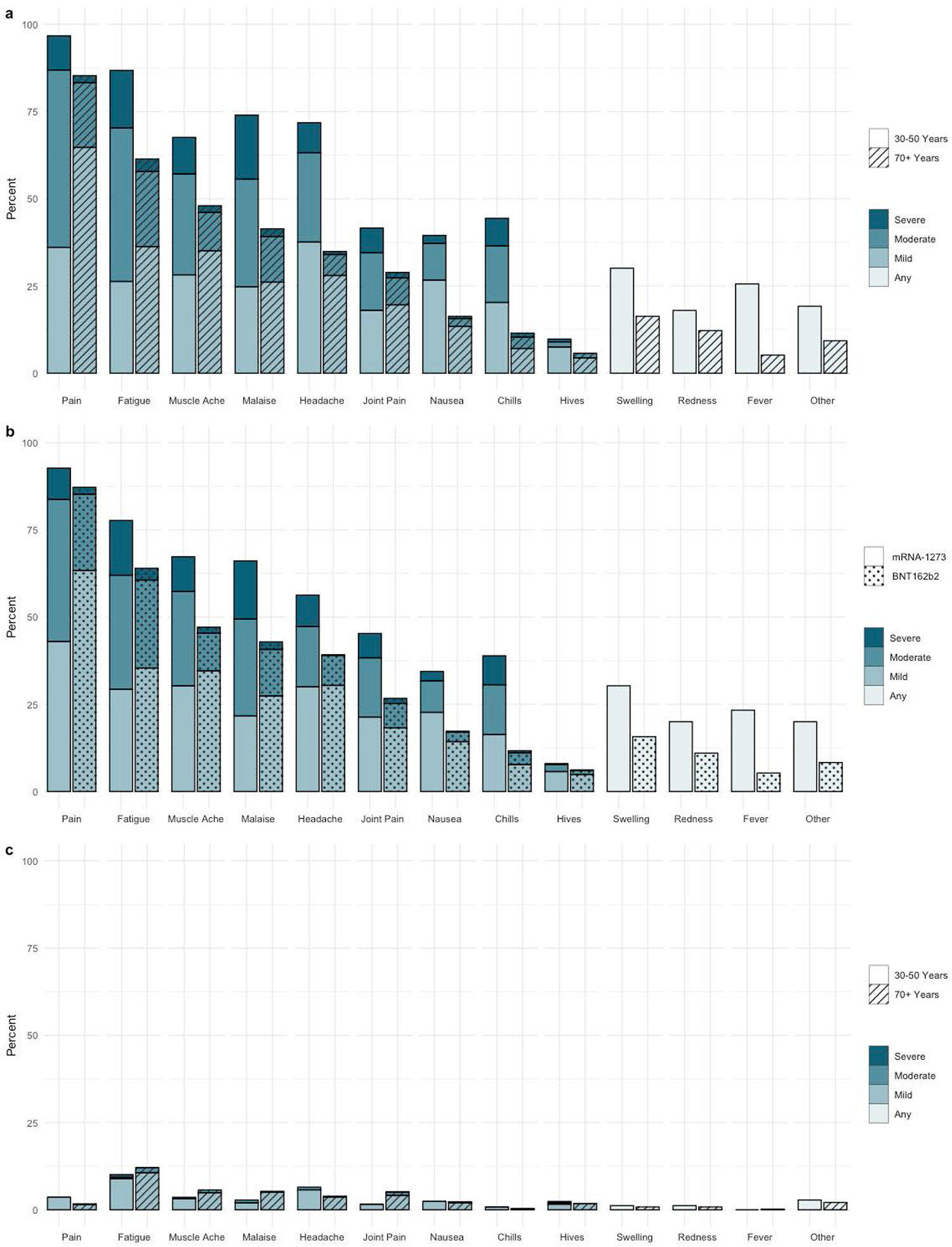
Self Reported Adverse events to Second vaccine dose a) Adverse events and reported severity during first 7 days post dose 2 by age cohort (n=955) b) Adverse Events and reported severity during first 7 days post dose 2 by vaccine type (n=938) c) Adverse Events and severity reported on day 7 post dose 2 by age cohort (n=905)

### Serology

The relative antibody ratios are available for 14 (1%) participants prior to their first vaccine, 83 (7%) three weeks post first vaccine, 966 (81%) prior to the second vaccine, 1002 (84%) two weeks after the second vaccine and 878 (74%) 12 weeks after the second vaccine. Figure 2 displays the violin plots of anti-spike and anti-RBD antibody ratios by time and age cohort. Detailed normalized antibody ratios by time and age are provided in (Supplemental Table 1).

**Figure 2:**
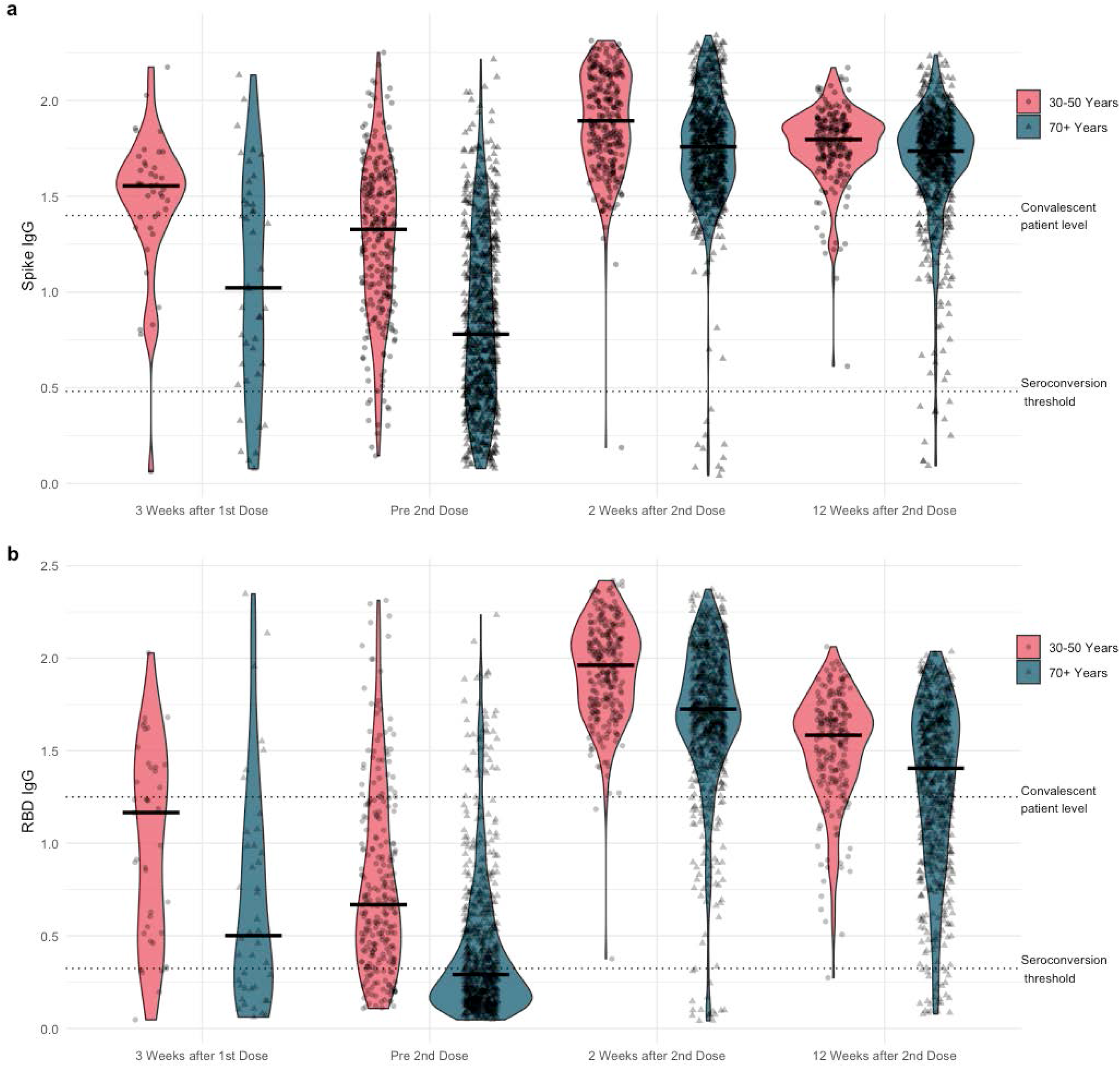
Violin plots of a) anti-Spike and b) anti-RBD (receptor binding domain) IgG normalized ratios by Time and by Age Cohort

Prior to the second vaccine dose, 25 (2·6%) participants had positive anti-NP indicating possible prior natural infection. Of these, 7 reported a prior COVID infection. An additional ten participants had their first positive anti-NP two weeks after their second dose and 4 twelve weeks after the second dose; none of these participants reported a COVID diagnosis on their monthly check-ins.

In the older cohort, the proportion with positive spike IgG antibodies increased from 518 (73%) prior to the second vaccine dose to 723 (99%) two weeks after the second dose. The proportion with positive RBD IgG antibody increased from 321 (46%) to 718 (98%) after the second vaccine dose. Of the 16 (2%) older participants negative for anti-RBD IgG two weeks after the second dose, 11 were negative for anti-spike IgG. Most were women (n=11), had cardiovascular disease (n=10), received two doses of BNT162b2 (n=11), and had their doses more than 8 weeks apart (n=12). There was an increase in median antibody ratios from before the second vaccine dose to two weeks after the second dose. At 12 weeks post second vaccine dose, the percentage with positive RBD antibodies remained high (96%), but the median antibody ratio decreased (Figure 2b).

The proportion of younger participants with positive antibody ratios increased from before the second vaccine dose to two weeks after the second dose; the proportion with positive spike IgG increased from 96% to 100% and RBD IgG increased from 84% to 100%. Younger participants also demonstrated an increase in median antibody ratios from before the second vaccine dose to two weeks after the second dose. Twelve weeks after the second dose, all had positive IgG antibodies to spike and all but one positive for RBD IgG, but similar to the older cohort, median RBD ratios decreased more than spike ratios (Figure 2).

Table 2 demonstrates results by brand. For both the younger (p<·006) and older cohort (p<·001), at two weeks and 12 weeks post second dose, RBD antibody ratios were higher for those receiving two doses of mRNA-1273 or one dose of mRNA-1273 and one dose of BNT162b2 and lower for those receiving two doses of BNT162b2 or other vaccine combinations.

**Table 2:**
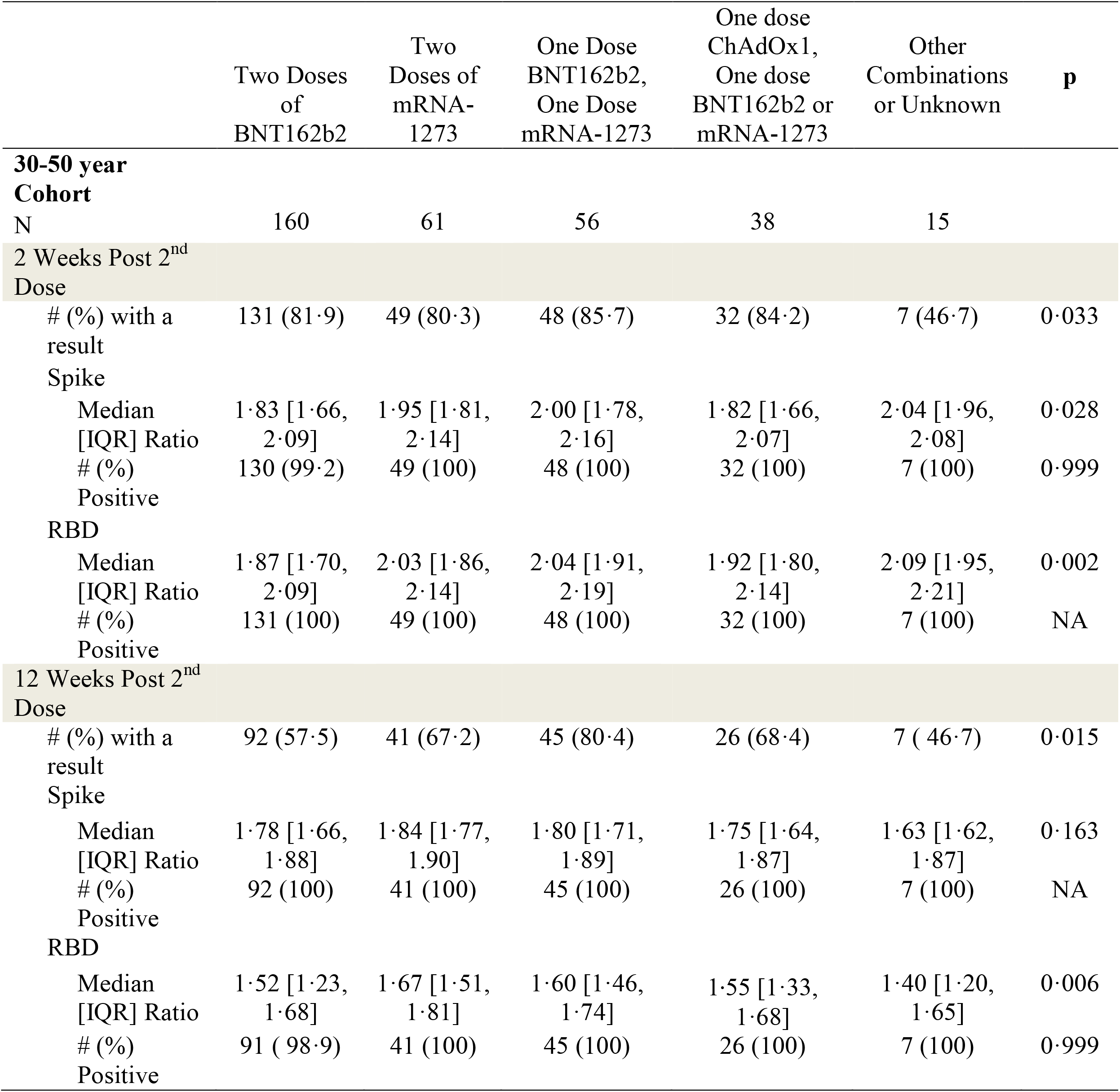
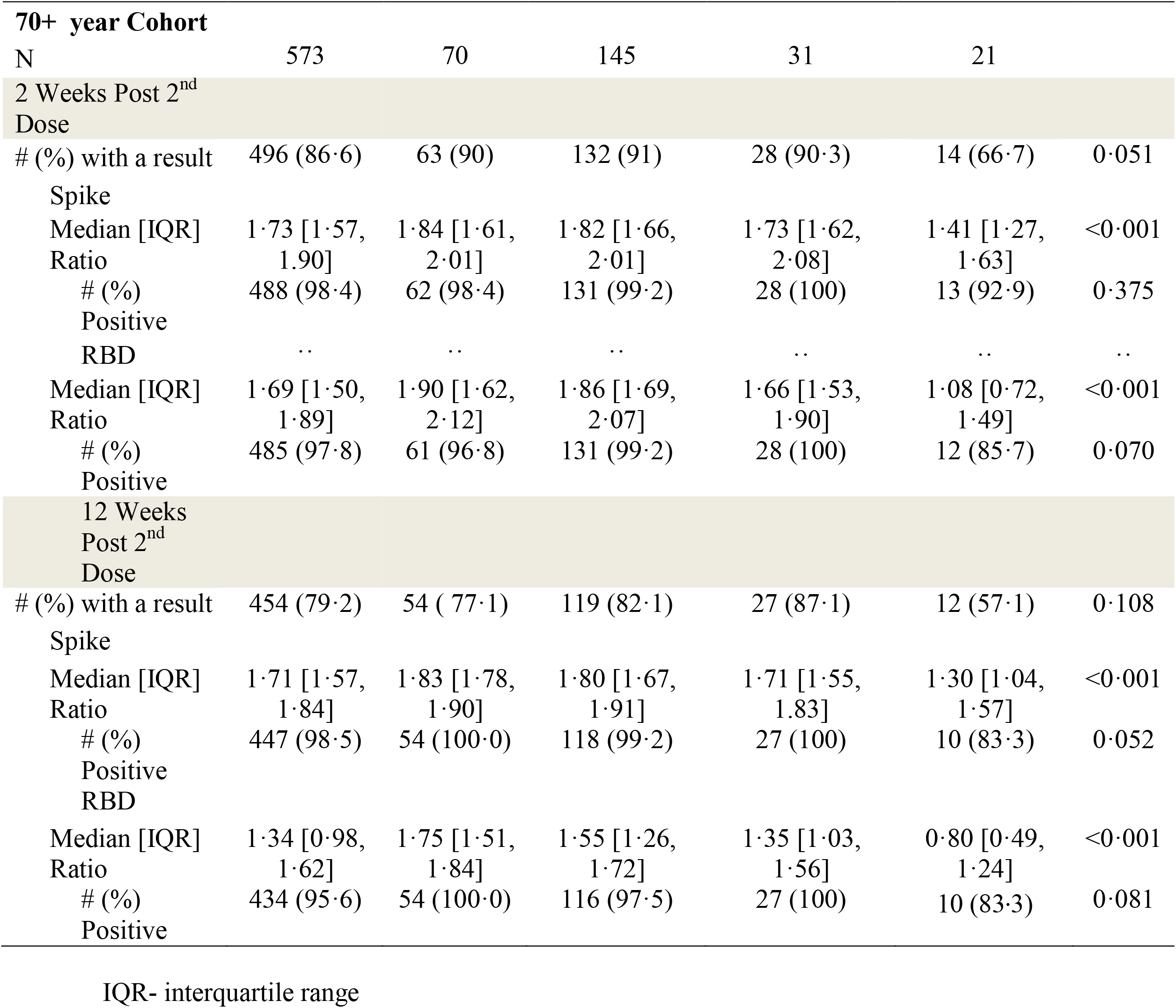
IgG antibody normalized ratios to spike and RBD (receptor binding domain) by Time and Vaccine Brand

In both cohorts, (Supplemental Figure 3) longer time between vaccine doses was associated with lower ratios of RBD IgG (PCC=-0·29, p<0·0001 for 30-50; PCC=-0·27, p<0·0001 for 70+) antibodies prior to receiving the second dose. This effect remained significant after the second dose although less pronounced and no longer significant at 12 weeks post second vaccine.

Table 3 is the univariable and multivariable linear regression models of positive antibody ratios to RBD IgG at 2 and 12 weeks post second vaccine dose. After adjusting for covariates, older participants had lower RBD ratios two weeks after the second dose compared to younger participants (p<·0001). A cancer diagnosis (p<·04) and a longer interval between doses (p<·001) was associated with lower anti-RBD ratios. Female or non-binary gender (p=·02), higher BMI (p=·05), and two doses of mRNA-1273 (p <·001) or one dose of each brand of mRNA vaccine (p<·001) (compared to two doses of BNT162b2) were associated with higher ratios of anti-RBD IgG. When using a 0·156 ul/well dilution, cardiovascular disease and immunosuppression were also associated with lower anti-RBD levels (Supplemental Table 2). At 12 weeks after the second vaccine dose, older participants had lower ratios of anti-RBD after adjusting for covariates (p<·01). Participants with two doses of mRNA-1273 or one dose of each brand of mRNA vaccine had higher anti-RBD ratios compared to those who received two doses of BNT162b2 p<·0001) as did female/non-binary participants (p<·01). Comorbidities, BMI, and dose interval were not associated with anti-RBD ratios at 12-weeks post second vaccine dose.

**Table 3:**
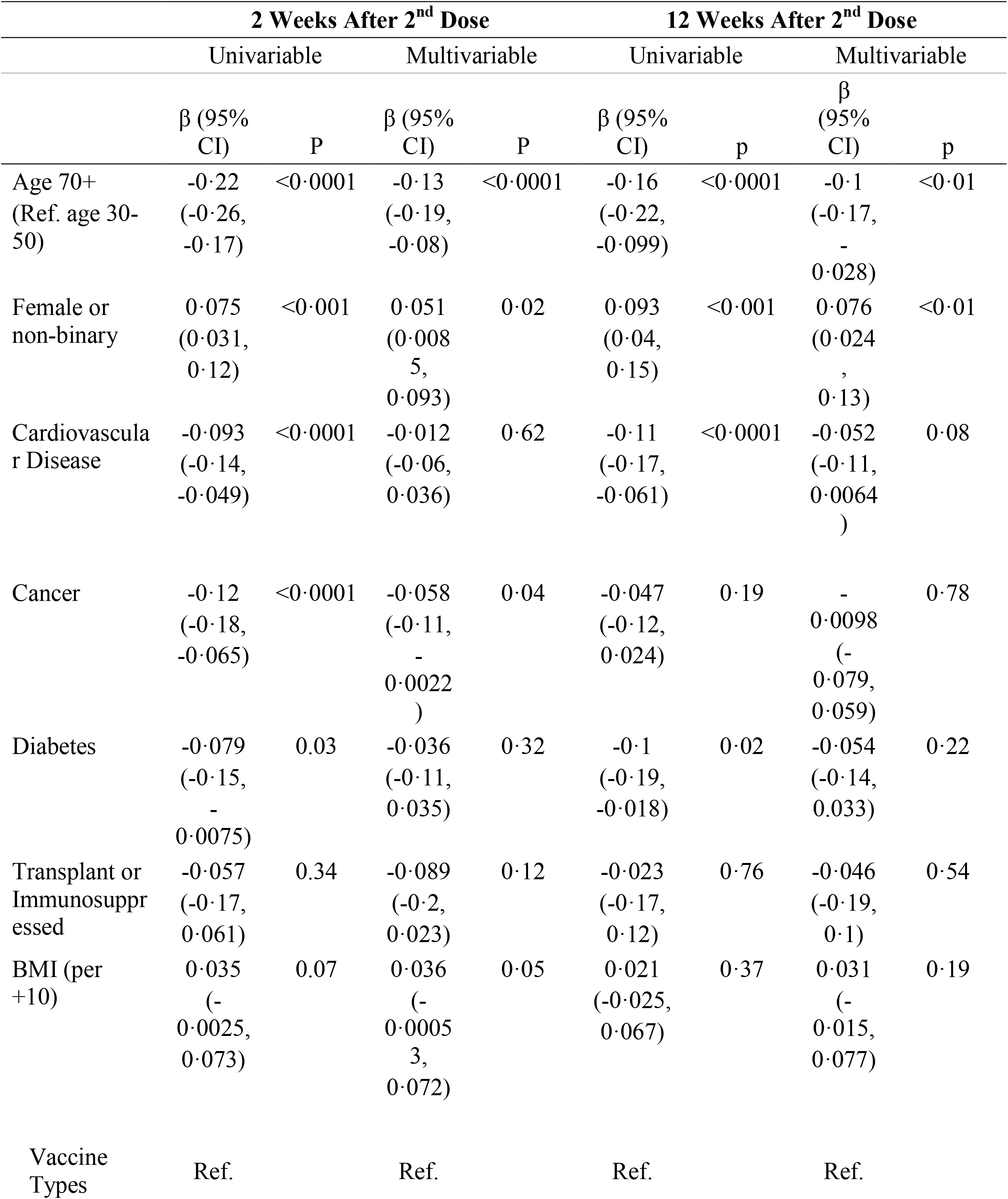
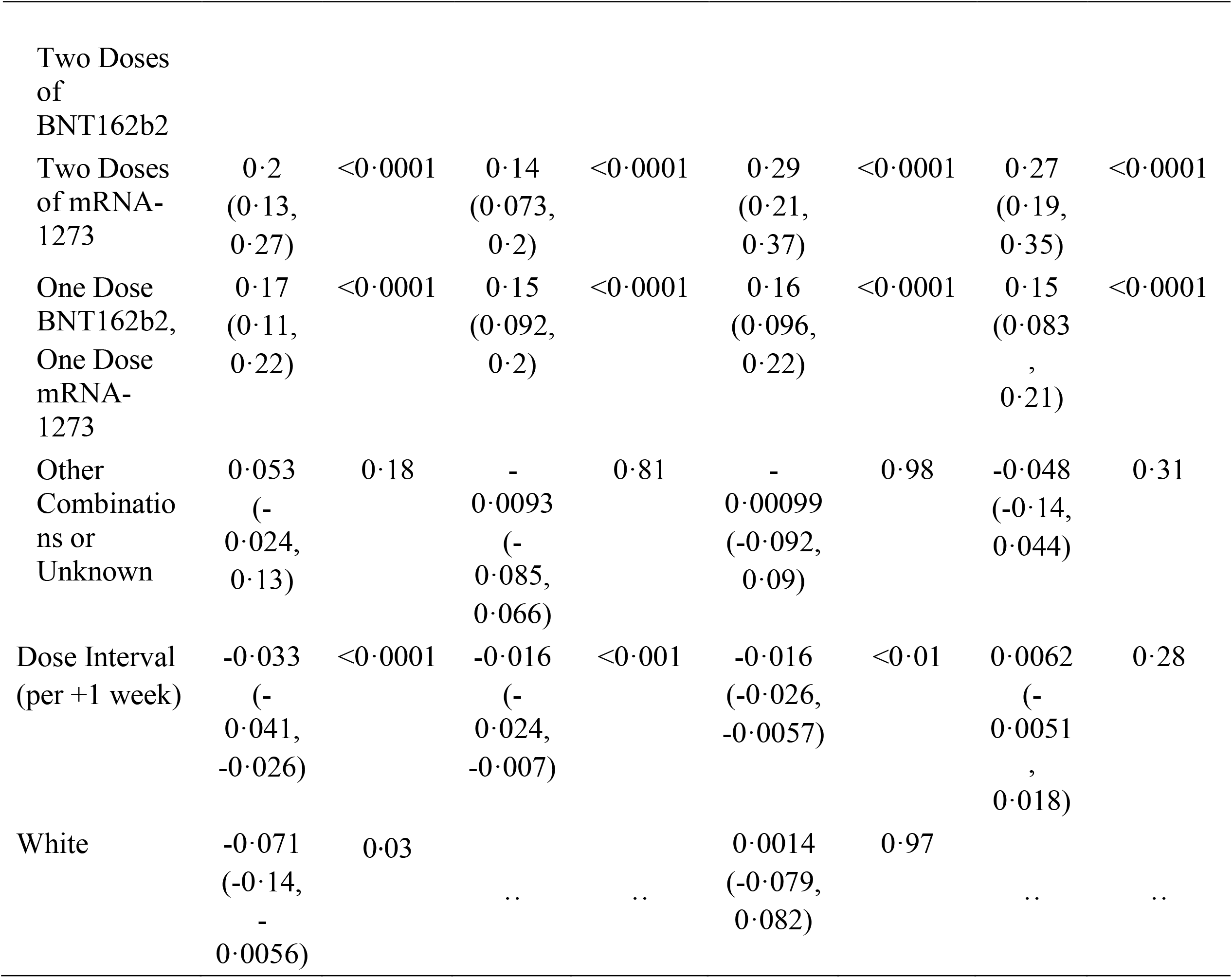
Linear Regression Models of Normalized RBD (receptor binding domain) IgG Antibody Ratios

## Discussion

We report the real time antibody response in the largest published cohort of community dwelling elders relative to a younger community cohort. Our older cohort (>70 years of age) had a less robust antibody response to COVID-19 vaccines than the younger cohort (aged 30-50 years). Although 98% of older and 100% of younger participants had a positive antibody response (normalized ratios of IgG to both RBD and spike above threshold) after two COVID vaccine doses, only 84% of the younger cohort and 46% of the older cohort had a positive antibody response prior to the second dose. Although successful in partially immunizing more of a population in a situation of limited supply with a single dose, our data reinforces the importance of receiving the two dose vaccination series.

Antibody to RBD is thought to most closely reflect neutralizing antibody ^[8, 13]^ at the population level and in our study, the normalized ratio of IgG antibody to RBD at two weeks post the second vaccine dose was higher in the younger than the older cohort with the peak normalized ratio of antibody similar to the median relative ratio of values of convalescent sera. At week 12 post second vaccine dose there was a significant decline in RBD IgG antibody ratio in both cohorts but remained higher for the younger cohort. Our data has demonstrated that age is an important predictor of antibody response irrespective of other demographic variables.

There has been considerable concern by the public as to which vaccine brand to receive and whether or not protection would be adequate with brand mixing. In our study at two weeks post second vaccine dose, those who received two doses of mRNA-1273 vaccine had higher antibody ratios than those receiving two doses of BNT162b2^[11, 14]^. The ratio for those receiving at least one dose of mRNA-1273 combined with a different brand was intermediate in both age cohorts. This is consistent with others who have demonstrated comparable antibody responses with brand mixing.^[15]^ A Veterans administration study provided clinical evidence that mRNA-1273 vaccine provides better protection from infection among older adults.^[16]^ The larger quantity of antigen in the mRNA-1273 vaccine is likely the explanation for these observations.

The optimal dose interval between vaccine doses is unclear. We showed a lower antibody response for each increasing week between doses both prior to and two weeks after the second vaccine dose but this effect was no longer significant at 12 weeks after the second dose after adjusting for other covariates. As only 4% of our participants received two doses within the three to four weeks recommended interval, we cannot determine whether delay beyond four weeks had any benefit as shown by others.^[17]^ Although additional reports have demonstrated higher serologic outcomes with longer dose intervals ^[18] [9]^ the outcomes may reflect difference in sampling time. In all studies the interval is not random and unmeasured confounders may influence antibody responses. Given the low seroconversion rate of our older cohort after the first dose, we recommend the second dose should not be delayed.

In other infection/vaccine studies such as influenza^[19]^, older adults and compromised persons have a less robust response, likely as a consequence of immunosenescence, leading to changes in vaccine strategies. This may also be necessary with COVID-19 vaccines^[20]^. In our study, in addition to age, gender, cardiovascular disease, lower weight, prior cancer, and immunosuppression were associated with lower antibody response. Other studies have shown lower antibody^[21]^ and neutralizing antibody responses and faster antibody level declines in older adults ^[21]^ and adults in long term care^[11]^ compared to staff^[22]^. A modelling study from the United Kingdom predicts a lower response in older participants and those with long-term health conditions.^[23]^ Consistent with these observations, vaccine effectiveness against COVID-19 hospitalization declined over time in a study of persons > 65 years ^[24]^. Understanding whether a specific antibody level has predictive power for vaccine efficacy at the individual level and what threshold confers a desired level of protection in different populations will help inform timing of further doses^[6, 25]^. Collectively age and comorbidity contribute to a less robust vaccine antibody response supporting the prioritization of booster doses in these populations. The elderly in long term care are anticipated to have more comorbidity and lower antibody responses than the elderly in the community, but direct comparison studies have not been completed. It may be necessary to have other strategies to overcome possible age-related limitations to COVID-vaccination such as the use of adjuvents, increased amounts or multiple doses, or more frequent boosters.

In our study the vaccines were safe and well tolerated. Although 95% of participants reported at least one adverse event, they tended to be mild and short lived. Adverse events were reported more commonly in our younger cohort and those receiving mRNA-1273. Our rates appeared to be higher than that previously reported ^[26-28]^, which may reflect real time symptom recording or the misinterpretation of symptoms related to underlying disease. However, by seven days after the second dose few participants reported residual symptoms.

Our unique and nimble study included electronic consent, questionnaires, symptom diaries and serial self-collected specimens. Our decentralized protocol and recruitment strategy enabled province-wide enrollment, including smaller communities without access to hospital-based research centers. The cohort is less diverse than planned reflecting the need for English and grasp of web-based technology. Additional strengths include the cohort size and retention and our ability to quickly adapt to changing vaccine dose and interval recommendations. Other population-based serology studies evaluating vaccine responses through DBS^[29, 30]^ have small population sizes, infrequent testing and younger individuals. Returning individual and group results through the platform kept our population engaged which will contribute to robust long term evaluation of the serology and the impact of the booster doses. Use of DBS enabled us to collect specimens at home but limits our ability to study the multifaceted immune response that importantly includes memory B-cells and T-cell responses.

Despite the challenges of rapid implementation of a digital platform, less familiarity of our target population with electronic platforms, tight and changing vaccine distribution timelines and the pressures of rapid kit distribution and postal services, we have successfully engaged and maintained a large cohort of older dedicated participants.

## Conclusion

We report on a successful large decentralized research program to study the IgG antibody response to COVID-19 vaccines. Our work provides data on the age-dependent limitations of antibody responses elicited after the first and second dose of COVID-vaccines, a vulnerable group that was underrepresented in the vaccine clinical trials. We demonstrated less robust responses with BNT162b2 relative to mRNA-1273 but no harm by extending dosing intervals. Large prospective population data will provide insight into future vaccine strategies for older adults as the correlates of protective immunity are increasingly understood.

## Data Availability

All data produced in the present study are available upon reasonable request to the authors.

## Acknowledgements

We acknowledge the technical support of the Network Biology Collaborative Centre and members of the Gingras lab for performing ELISA analysis.

Antigens, protein standards and secondary antibodies for ELISA were kindly provided by The Pandemic Response Challenge Program of the National Research Council of Canada (Dr. Yves Durocher); positive and negative control samples for ELISA assay calibration were from the National Microbiology Laboratory, Public Health Agency of Canada (Dr. John Kim). The robotics equipment used is housed in the Network Biology Collaborative Centre at the Lunenfeld-Tanenbaum Research Institute, a facility supported by the Canada Foundation for Innovation, the Ontario Government, and Genome Canada and Ontario Genomics (OGI-139).

Anne-Claude Gingras leads the Functional Genomics and Structure-Function Pillar of CoVaRR-Net and is the Canada Research Chair (Tier 1) in Functional proteomics. Dr. Rochon is the RTOERO Chair in Geriatric Medicine at the University of Toronto.

## Author contributions

SW is the principal investigator and takes overall responsibility for the integrity of the work from inception to publication.

LS, JR are the statisticians who developed the statistical plan and completed the analysis.

BW was responsible for study conception and developing the data sharing agreement with the Ministry of Ontario.

RC, RR were the main study coordinators responsible for ethics submission, day to day study conduct and data collection and cleaning.

AM, PR, CG, AO were involved in study conception and critical review of study design and manuscript.

KC, RD, AP conducted the lab analysis of the DBS under the supervision of ACG who also developed the laboratory techniques.

AS, DM, JS, LP developed the digital platform and modifications, and helped with data collection and cleaning under the supervision of MB.

SW, LS had full access to all data in the study and take responsibility for the integrity of the data and the accuracy of the data analysis.

All authors reviewed and revised the manuscript drafts and final version. All authors agree to be accountable for the accuracy and integrity of the part of the work they have participated in.

## Data Sharing Statement

The study funder, the COVID-19 Immunology Task Force (CITF), has a data sharing protocol for all funded projects. We will transfer relevant anonymized study data as available to the CITF as a part of these standard data sharing requirements. This is submitted together with a data dictionary defining each field in the set. External researchers will be able to submit a request to the CITF to receive access to all CITF data through their data access committee. The CITF will employ a rigorous checklist to ensure that these external requests follow all necessary ethical and privacy protocols.

The data provided to the CITF will be stored on the CITF Database. The data on the CITF Database will be held under the custodianship of McGill University or one of its collaborators and be shared via the cloud, both nationally and internationally. Data in the CITF Database can be used by researchers across Canada and in other countries following Data Access Committee (DAC) approval. These transfers will also be made in compliance with Canadian law and research ethics.

A DAC will be responsible for reviewing applications for access to the data and for approving applications that respect the privacy and access policies of the CITF. The DAC will require that researchers confirm that their intended research activities have received necessary ethics approvals. The data may also be shared with other COVID-19 research databases that follow similar protections and procedures as the CITF Database.

Further the study protocol, statistical analysis plan, informed consent form and full protocol are available on the study website www.stopcov.ca

