## Supplementry materials for "Safety and Efficacy of Preventative COVID Vaccines: The StopCoV Study"

Supplementary Tables:

s Table 1: Normalized IgG antibody Ratios to Spike and Receptor binding domain (RBD) by Time from Vaccine dose and Age Cohort

| | **30-50 years** | **70+ years** | **P value** |
|---|---|---|---|
| N | 340 | 853 | |
| 3 Weeks Post 1st Dose | | | |
| # (%) with a result | 41 ( 12.1) | 42 ( 4.9) | <0.001 |
| Spike | | | |
| Median [IQR] Ratio | 1.55 [1.39, 1.66] | 1.02 [0.62, 1.47] | <0.001 |
| # (%) Positive | 40 ( 97.6) | 35 (83.3) | 0.068 |
| RBD | | | |
| Median [IQR] Ratio | 1.17 [0.52, 1.41] | 0.50 [0.22, 0.99] | 0.003 |
| # (%) Positive | 35 ( 85.4) | 26 (61.9) | 0.030 |
| Pre 2nd Dose | | | |
| # (%) with a result | 260 ( 76.5) | 706 (82.8) | 0.016 |
| Spike | | | |
| Median [IQR] Ratio | 1.33 [0.99, 1.58] | 0.78 [0.47, 1.19] | <0.001 |
| # (%) Positive | 249 ( 95.8) | 518 (73.4) | <0.001 |

| | 30-50 years | 70+ years | P value |
|---|---|---|---|
| RBD | | | |
| Median [IQR] Level | 0.67 [0.41, 1.12] | 0.29 [0.16, 0.54] | <0.001 |
| # (%) Positive | 217 ( 83.5) | 321 (45.5) | <0.001 |
| 2 Weeks Post 2nd Dose | | | |
| # (%) with a result | 268 ( 78.8) | 734 (86.0) | 0.003 |
| Spike | | | |
| Median [IQR] Ratio | 1.89 [1.70, 2.12] | 1.76 [1.58, 1.93] | <0.001 |
| # (%) Positive | 267 ( 99.6) | 723 (98.5) | 0.262 |
| RBD | | | |
| Median [IQR] Ratio | 1.96 [1.75, 2.13] | 1.73 [1.54, 1.96] | <0.001 |
| # (%) Positive | 268 (100.0) | 718 (97.8) | 0.031 |
| 12 Weeks Post 2nd Dose | | | |
| # (%) with a result | 211 ( 62.1) | 667 (78.2) | <0.001 |
| Spike | | | |
| Median [IQR] Ratio | 1.80 [1.70, 1.89] | 1.74 [1.60, 1.86] | <0.001 |
| # (%) Positive | 211 (100.0) | 657 (98.5) | 0.157 |
| RBD | | | |
| Median [IQR] Ratio | 1.58 [1.36, 1.71] | 1.41 [1.03, 1.66] | <0.001 |
| # (%) Positive | 210 ( 99.5) | 642 (96.3) | 0.027 |

s Table 2: Linear Regression Models of normalized IgG RBD (receptor binding domain) Antibody Ratios 2 Weeks After 2nd Dose (0.156 Dilution)

| | Univariable | | Multivariable | |
|---|---|---|---|---|
| | β (95% CI) | p | β (95% CI) | p |
| Age 70+ (Ref. age 30-50) | -0.28<br>(-0.33, -0.22) | <0.0001 | -0.16<br>(-0.22, -0.094) | <0.0001 |
| Female or non-binary | 0.1<br>(0.052, 0.16) | <0.0001 | 0.076<br>(0.028, 0.12) | <0.01 |
| Cardiovascular Disease | -0.13<br>(-0.18, -0.081) | <0.0001 | -0.056<br>(-0.11, -0.002) | 0.04 |
| Cancer | -0.14<br>(-0.21, -0.071) | <0.0001 | -0.06<br>(-0.12, 0.0027) | 0.06 |
| Diabetes | -0.085<br>(-0.17, -0.00083) | 0.05 | -0.033<br>(-0.11, 0.047) | 0.42 |
| Transplant or Immunosuppressed | -0.13<br>(-0.27, 0.0088) | 0.07 | -0.17<br>(-0.29, -0.039) | 0.01 |
| BMI (per +10) | 0.11<br>(0.064, 0.15) | <0.0001 | 0.12<br>(0.074, 0.16) | <0.0001 |
| Vaccine Types | | | | |

| | Univariable | | Multivariable | |
|---|---|---|---|---|
| | β (95% CI) | p | β (95% CI) | p |
| Two Doses of BNT162b2 | Ref. | | Ref. | |
| Two Doses of mRNA-1273 | 0.36 (0.28, 0.44) | <0.0001 | 0.29 (0.21, 0.36) | <0.0001 |
| One Dose BNT162b2, One Dose mRNA-1273 | 0.22 (0.16, 0.29) | <0.0001 | 0.19 (0.13, 0.25) | <0.0001 |
| Other Combinations or Unknown | 0.068 (-0.021, 0.16) | 0.13 | -0.015 (-0.1, 0.07) | 0.73 |
| Dose Interval (per +1 week) | -0.042 (-0.051, -0.032) | <0.0001 | -0.017 (-0.026, -0.007) | <0.001 |
| White | -0.12 (-0.2, -0.043) | <0.01 | | |

BMI: body mass index

s Figure 3 Legend

Normalized IgG Ratios to RBD (receptor binding domain) pre and 2 weeks post second vaccine dose by time interval between doses

a) Pre- second dose- anti-RBD IgG

b) 2 weeks post second dose- anti RBD- IgG

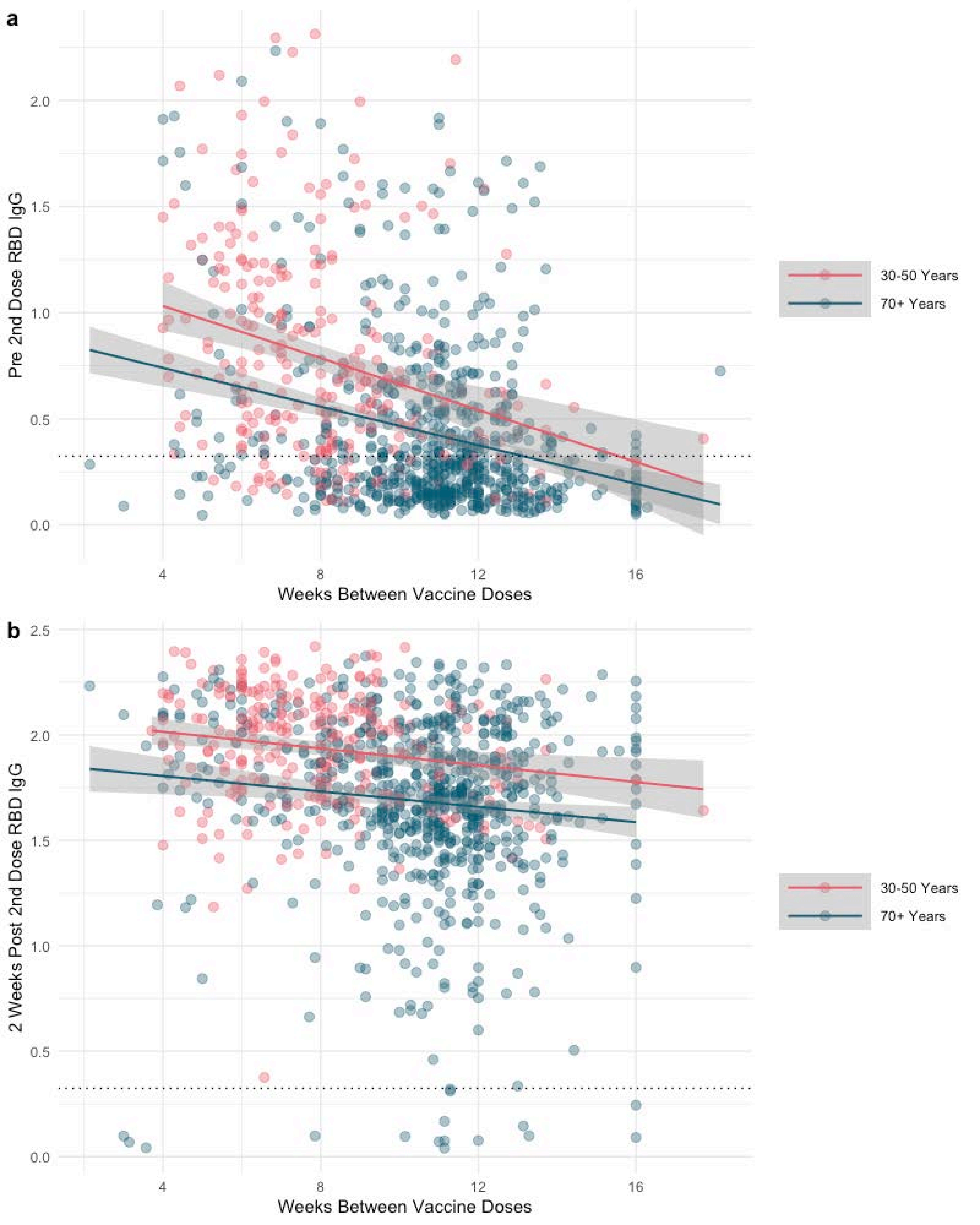
a
Pre 2nd Dose RBD IgG
0.0
0.5
1.0
1.5
2.0
4
8
12
16
Weeks Between Vaccine Doses
30-50 Years
70+ Years
b
2 Weeks Post 2nd Dose RBD IgG
0.0
0.5
1.0
1.5
2.0
2.5
4
8
12
16
Weeks Between Vaccine Doses
30-50 Years
70+ Years
